# Impaired flexible reward learning is associated with blunted reinforcement sensitivity and attenuated learning and choice signals in ventral striatum and parietal cortex of ADHD patients

**DOI:** 10.1101/2023.04.14.23288555

**Authors:** Hans-Christoph Aster, Maria Waltmann, Anika Busch, Marcel Romanos, Matthias Gamer, Betteke Maria van Noort, Anne Beck, Viola Kappel, Lorenz Deserno

## Abstract

Reward-based learning and decision-making are prime candidates to understand symptoms of attention deficit hyperactivity disorder (ADHD). However, only limited evidence is available regarding the neurocomputational underpinnings of the alterations seen in ADHD. This particularly concerns the flexible behavioral adaption in dynamically changing environments, which is challenging for individuals with ADHD. One previous study points to elevated choice switching in adolescent ADHD, which was accompanied by disrupted learning signals in medial prefrontal cortex.

In the present study, we investigated young adults with ADHD (n=17, 18-32 years) and age and sex matched controls (n=17, 18-30 years) using a probabilistic reversal learning experiment during functional magnetic resonance imaging (fMRI). The task requires continuous learning to guide flexible behavioral adaptation to changing reward contingencies. To disentangle the neurocomputational underpinnings of the behavioral data, we used detailed reinforcement learning (RL) models, which informed the analysis of fMRI data.

ADHD patients performed worse than controls particularly in trials before reversals, i.e., when reward contingencies were stable. This pattern resulted from ‘noisy’ choice switching regardless of previous feedback. RL modelling showed decreased reinforcement sensitivity and enhanced learning rates for negative feedback in ADHD patients. At the neural level, this was reflected in diminished representation of choice probability in the left posterior parietal cortex in ADHD. Moreover, modelling showed a marginal reduction of learning about the unchosen option, which was paralleled by an equally marginal reduction in learning signals incorporating the unchosen option in the left ventral striatum.

Taken together, we show that flexible behavioral adaptation in the context of dynamically changing reward contingencies is impaired in ADHD. This is due to excessive choice switching (‘hyper-flexibility’), which can be detrimental or beneficial depending on the learning environment. Computationally, this results from blunted sensitivity to reinforcement. We detected neural correlates of this blunted sensitivity to reinforcement in the attention-control network, specifically in the parietal cortex. These neurocomputational findings are promising but remain preliminary due to the relatively small sample size.

## Introduction

Attention deficit hyperactivity disorder (ADHD), a common child and adolescent psychiatric disorder (Faraone, Asherson et al. 2015), is characterized by its core symptoms of hyperactivity, inattention, and impulsivity. Reward-based learning and decision-making are prime candidates that may underlie symptoms, as alterations were reported in some of these domains (Mowinckel, Pedersen et al. 2015, Marx, Hacker et al. 2021). However, only limited evidence is available regarding the neurocomputational underpinnings of reward learning and decision-making in ADHD. This is particularly true with respect to flexible behavioral adaption in dynamically changing environments, which may be challenging for individuals with ADHD (Humphreys, Tottenham et al. 2018) due to attentional and learning deficits.

How individuals learn from positive and negative reward feedback and guide decisions accordingly can be formalized by computational models of reinforcement learning (Sutton and Barto 1998). At the core of RL models are reward prediction errors (RPEs), which reflect the differences between delivered and expected reward. Neurally, prediction errors are signaled by phasic release of midbrain dopamine (Hollerman and Schultz 1998, Schultz 2013), with corresponding echoes of neural activity in the striatum as well as other brain regions (Pine, Sadeh et al. 2018). Human functional neuroimaging studies reported correlates of RPEs in the midbrain, striatum and several cortical regions (O’Doherty, Dayan et al. 2004, D’Ardenne, McClure et al. 2008, Daw, Gershman et al. 2011, Deserno, Huys et al. 2015). Individual differences in neurobehavioral correlates of RL have been indeed linked to a variety of dopamine measures available in humans, including pharmacological manipulations (Pessiglione, Seymour et al. 2006, Westbrook, van den Bosch et al. 2020, Deserno, Moran et al. 2021), neurochemical positron emission tomography (PET) (Deserno, Huys et al. 2015, Westbrook, van den Bosch et al. 2020, Calabro, Montez et al. 2023) and specific genotypes (Frank, Moustafa et al. 2007, Dreher, Kohn et al. 2009).

In patients with ADHD, neurochemical studies reported altered dopamine neurotransmission and presumably lower baseline dopamine levels (Fusar-Poli, Rubia et al. 2012). Brain activation, measured with fMRI during the anticipation and delivery of rewards, was reported to be disrupted (Plichta and Scheres 2014, von Rhein, Cools et al. 2015), in particular in the ventral striatum during reward anticipation (Plichta and Scheres 2014). This line of work supports hypothetical alterations in RL and its neural underpinnings. However, evidence based on studies that directly test learning and apply computational modeling (Ziegler, Pedersen et al. 2016, Véronneau-Veilleux, Robaey et al. 2022) is missing. A particular challenging scenario for individuals with ADHD is not only to learn from reward to guide decision-making but also to strike a balance between exploration and exploitation when action-outcome contingencies change dynamically. This capacity can be examined using reversal learning (Reiter, Heinze et al. 2017, Waltmann, Herzog et al. 2023). Reinforcement learning has been shown to undergo substantial neurodevelopmental changes (Nussenbaum and Hartley 2019, Weiss, Kruppa et al. 2021, Scholz, Waltmann et al. 2023, Waltmann, Herzog et al. 2023), and has been used to study a wide range of psychiatric disorders (Chantiluke, Barrett et al. 2015, Geisler, Ritschel et al. 2017, Reiter, Heinze et al. 2017). Yet, there is only one study available that directly examined RL in adolescent ADHD patients during fMRI (Hauser, Iannaccone et al. 2014). This study revealed noisy switching behavior in ADHD patients, which may computationally arise from enhanced levels of decision noise, an impairment in distinctly representing values of alternative choice options. In the study by Hauser et al. (2014) this was accompanied by reduced activation to RPEs in the medial prefrontal cortex. Our study aimed to extend these findings by investigating RL in adult ADHD patients. Furthermore, this study explored for the first time explicitly whether impaired learning of the selected action or impaired simultaneous learning of the unselected action caused the difficulties in RL.

In n=17 patients and n=17 controls, we closely followed the initial study by Hauser et al. (2014) by examining reversal learning during fMRI with extended RL modelling and more detailed computational fMRI analysis. We hypothesized that altered task performance would be driven by noisy choice switching, computationally accounted for by enhanced decision noise. In our RL models, we addressed not only learning from the chosen option (single-update learning) but also learning from the option that was not chosen (double-updating). Thus, we explored whether differences in these types of learning also contributed to the observed behavioral alterations seen in ADHD. On the neural level, we dissociated correlates of RPE with respect to single- and double-update learning. We further analyzed the neural correlates of choice probability, which closely reflects decision noise. We focused these analyses on the ventromedial prefrontal cortex and the ventral striatum, which were previously reported to be altered in ADHD, and where we expected reduced correlates of RPEs and choice probability.

## Methods

### Study protocol

Before participation, all participants provided written informed consent. Ethical approval was obtained through the ethics committee of the German Psychological Society (DGPs registration number: HSAAS04082008DGPS). Data were collected between 2008-2011.

All participants completed several diagnostic and neuropsychological assessments before the MRI acquisition. All patients fulfilled the DSM-IV-TR criteria for ADHD combined subtype as assessed by clinical experts with the structured assessment scale ‘ADHD Diagnostic Checklist’ (Rösler, Retz et al. 2005). ADHD symptomatology in childhood was assessed retrospectively with the ‘Wender Utah Rating Scale – German short form’ (WURS, (Retz-Junginger, Retz et al. 2002)). The current severity of ADHD symptomatology was examined via the ‘Conner’s Adult ADHD Rating Scale’ (CAARS, (Christiansen, Hirsch et al. 2013)). To exclude the presence of other Axis I or Axis II disorders, subjects were interviewed using the SCID-I and –II (Wittchen and Pfister 1997). Furthermore, the specific presence of substance abuse, amongst others due to its role in reward processing, was examined via the Composite International Diagnostic Interview (Wittchen and Pfister 1997). To rule out previous or current ADHD symptoms or other psychiatric disorders in the control group, we used the same diagnostic assessments. Finally, handedness was assessed via the ‘Handedness Questionnaire’ (Coren 1993). Forty-eight hours before the study appointment, the ADHD patients discontinued their intake of psychostimulants.

The neuropsychological assessment consisted of a language-independent measure for IQ, the Culture Fair Test (CFT-20-R (Weiß and Weiß 2008)), a Digit Span task (Von Aster, Neubauer et al. 2006) and the Trail-Making-Test (TMT) Part A and B (Reitan 1958). The Digit Span task measures verbal working memory capacity. It consists of two conditions: forward and backward. A span of 6-7 is considered an average score. The TMT assesses visual attention and processing speed in Part A and executive control and flexibility in Part B. The TMT Part B and A difference score provides a more precise measure of task-switching ability. Depending on the homogeneity of variances and normal distribution of the data, the neuropsychological data were compared between groups via independent samples t-tests, Mann-Whitney-U tests or Welch tests using the Jasp Toolbox (JASP Team (2022). JASP (Version 0.16.4) [Computer software]. Alpha was set at 0.05.

Exclusion criteria for HCs were 1) left-handedness, 2) current psychiatric diagnosis according to ICD-10 or DSM-IV-TR, except alcohol abuse, 3) the presence of neurological disorders, 4) a first-degree family member suffering from a neurological or psychiatric disorder or 5) currently taking psychotropic medication. HCs were recruited via advertisements in the community.

### Reversal learning paradigm

During functional MRI (fMRI) acquisition, participants performed a reversal learning task (Schlagenhauf, Rapp et al. 2013, Schlagenhauf, Huys et al. 2014, Deserno, Beck et al. 2015). The task required participants to choose between one of two geometric figures with different reward probabilities. After each choice, they received positive (green smiley) or negative feedback (red frowning face). The chosen stimulus and the feedback remained visible for 1 second. If participants did not choose a target within 2 seconds, the trial was rated as incorrect. A fixation cross was shown between the trials. The interval had a varying duration of 1 to 6.5 seconds (exponentially distributed). The task consisted of two runs of 100 trials each with a short break in between. During each run, the participants were exposed to three types of blocks in which the reward probabilities for a correct choice (right figure vs. left figure) were 80:20, 20:80 and 50:50. The block changed based on performance, i.e., after a minimum of 10 subsequent trials if participants reached 70% correct choices, or automatically after a maximum of 16 trials. Thus, each block was encountered between twice and four times during each run. Reversal learning could only take place between the 80:20 and 20:80 blocks because learning is not possible in 50:50 conditions. Hence, only these block types were included in the initial analysis of choice behavior.

### Analysis of behavorial data

Trials with correct choices (regardless of whether positive feedback was actually received as a result of the 80/20 probabilistic), coded as 1 vs. 0, as well as trials with a different chosen response as in the previous trial (Switching, coded as 1 vs. 0), and the reaction time per trial with a valid response were extracted for each trial. The five trials after a reversal were considered as post-reversal phase, the remaining trials as pre-reversal. Generalized linear mixed models were used to analyze behavior:

1. [Correct choices ∼ Group * Phase + (1 + Group * Phase |subject)],
2. [Switching ∼ Group * Previous feedback + (1 + Group * Previous feedback |subject)],
3. [Reaction Time ∼ Group * Previous feedback + (1 + Group * Previous feedback |subject)].

We used a full random structure (random intercepts, random slopes, correlation of slopes) (Barr, Levy et al. 2013). Except for the model with reaction time as a dependent variable, a binomial family was chosen and the logit link function was used. In the reaction time model, an inverse Gaussian family was chosen and an identity link function was used, since it was shown that this fits reaction time data best (Lo and Andrews 2015).

### Computational modelling of reinforcement learning

We analyzed the behavioral data using Q-learning models of reinforcement learning (Watkins and Dayan 1992). Thus, for each model, we identified the parameters which best accounted for each individual’s observed history of choices and outcomes. Our initial model was a single-update model, which only updates (or learns) the value of the chosen action Q_a,t_: Q_a,t_ +1 = Q_a,t_ + α(r - Q_a,t_). This value is updated in each trial by the prediction error δ = r-Q_a,t_. The rate to which prediction errors influence the update of Q value is captured by the learning rate α. Because reward probabilities of the two available actions in the reversal learning task are perfectly anti-correlated, the feedback of the chosen action could also influence the Q-value of the non-chosen action. We therefore additionally defined a double-update model, which updates both actions simultaneously to opposite directions: Q_a(unchosen),t_ +1 = Q_a(unchosen),t_ + α((-r) - Q_a(unchosen),t_). It is conceivable that individuals vary in the degree of using double updating and thus we included a weighting parameter (K) that quantifies the degree of double-updating in some models: Q_a(unchosen),t_ +1 = Q_a(unchosen),t_ + κα((-r) - Q_a(unchosen),t_).

Further, since there could be inter-individual differences in the extent to which the current prediction error impacts updating depending on positive or negative feedback, different learning rates for wins and losses were implemented in some models (Q_a,t+1_ = Q_a,t_ + α_win/loss_ (r - Q_a,t_)).

Lastly, we examined decision noise, which is determined by the degree to which values of choice options are represented distinctly. Typically, this is determined by passing values to a sigmoid softmax function with an individually varying steepness parameter, which scales the differences between values and thus determines choice probabilities. Here, we used a slightly different but largely equivalent approach by implementing a reinforcement sensitivity parameter (ρ): Q_a,t_ +1 = Q_a,t_ + α(ρr - Q_a,t_). ρ is a free parameter that determines the maximum difference between values by defining the upper bound of the Q-values. These values scaled by ρ are then entered into a softmax function with steepness fixed to 1. While it was shown that this has an equivalent effect on choice probability as a steepness parameter, it is straightforward to implement differences in reinforcement sensitivity to positive and negative outcomes (Huys, Pizzagalli et al. 2013). Further, reinforcement sensitivities have improved estimation properties (Huys, Pizzagalli et al. 2013, Katahira 2015) and higher reliability (Waltmann, Schlagenhauf et al. 2022). Thus, in some of our models, reinforcement sensitivity was again distinguished for sensitivity to positive and negative feedback.

To summarize, by combining different learning rates and levels of reinforcement sensitivity, four models were created. These models were then estimated for single-update, double-update, and variable double-update scenarios, resulting in a total of 12 fitted models. To compare these models, the integrated Bayesian Information Criterion (iBIC) was used.

For hierarchical model estimation, we used the emfit toolbox in MATLAB R2020b (Huys 2017). Model estimation was performed to obtain maximum a posteriori estimation with empirical priors based on the trial-by-trial data of all participants. We have previously shown that this hierarchical estimation leads to improved reliability (Waltmann, Schlagenhauf et al. 2022). An expectation maximization procedure was used (Huys, Eshel et al. 2012). Since the model with only one reinforcement sensitivity can logically only assign positive values for the sensitivity, its value was transformed exponentially to ensure positive values. To keep the learning rate (α) and weighting parameter (κ) between 0 and 1, these parameters were inverse logit transformed.

The modeling parameters reinforcement sensitivity and learning rate (both for positive and negative feedback) were analyzed with linear mixed models using the Jasp Toolbox (JASP Team (2022). JASP (Version 0.16.4) [Computer software]. This resulted in two models with full random structure (1. Reinforcement sensitivity ∼ group * feedback + (1 + group * feedback |subject), 2. Learning rate ∼ group * feedback + (1 + group * feedback |subject)). The parameter kappa, which expresses the weighting between single and double updating, was compared between groups with a t-test after testing for normal distribution and equality of variance. Pearson correlation coefficients were calculated to discover possible associations between symptom expression and modeling parameters in an exploratory analysis. All variables were z-standardized before the correlation analysis.

### Functional MRI data acquisition

Imaging was conducted using a 3 Tesla GE Sigma Scanner with an eight channel head coil to acquire gradient echo T2*-weighted echo-planar images with blood oxygenation level-dependent (BOLD) contrast. Twenty-nine slices were acquired, covering the whole brain, with 4 mm thickness, 2 × 2 mm^2^ in-plane voxel resolution, repetition time (TR) = 2.3 ms, echo time (TE) = 27 ms and a flip angle α = 90°. T1-weighted structural images were acquired with TR = 7.8 ms, TE = 3.2 ms, α = 20°, matrix size = 256 × 256, slice thickness = 1 mm, voxel size = 1 × 1 × 1 mm. Right before the MRI acquisition, all participants were vigilant as assessed by the Stanford Sleepiness Scale (*M*_ADHD_= 2.12 ± 0.60, *M*_HC_ = 2.35 ± 0.70; *p* = .301, *d* = -0.35).

### Functional MRI data preprocessing

fMRI data were analysed with SPM8 (Wellcome Department of Imaging Neuroscience). ArtRepair was used to remove noise spikes and to repair bad slices within a particular scan and bad slices were repaired by interpolation between adjacent slices (Paul K Mazaika 2005). Data was then corrected for delay of slice time acquisition and was motion corrected using realignment. The images were then registered into the Montreal Neurological Institute (MNI) space by using the normalised parameters generated during the segmentation of each participant’s anatomical T1-image (Ashburner and Friston 2005). Spatial smoothing with an isotropic Gaussian kernel of 8 mm full width at half-maximum (FWHM) kernel was applied to the images.

### Model informed fMRI Analysis

In the first-level general linear model, onsets of feedback, cue and missing trials were convolved with the hemodynamic response function and the 6 motion parameters were added as regressors of no interest. As orthogonalized parametric modulators on the feedback regressor, we added, for each person, the trial-by-trial prediction errors (PEs) from the best fitting RL model. This included, first, the single update (SU) PEs from the best SU model and the double update (DU) PEs from the best-fitting model. Due to high collinearity between PEs and to isolate unique variance of the DU PEs, we subtracted SU PEs from DU PEs for each trial (Daw, Gershman et al. 2011). This approach has already been applied successfully in previous studies (Reiter, Deserno et al. 2016, Reiter, Heinze et al. 2017, Waltmann, Herzog et al. 2023). As orthogonalized parametric modulators to the cue onset, we added two model-derived trial-by-trial regressors. The choice probability maps the individual expected values of the choices per trial which are drawn from the best fitting DU model. The larger the difference in expected values between the two choices, the more likely an individual will choose one of the two options. From the choice probabilities, we constructed a regressor reflecting trial-by-trial model-fit, where choices predicted with below-chance accuracy (<50%) were coded as 1 (noisy or explorative behavior) and 0 otherwise. This regressor addresses brain activation associated with noisy or explorative behavior and removes variance solely associated with poor model fit (Waltmann, Herzog et al. 2023).

At the second level, a full factorial model was used on SU PEs and DU PEs with group and type of RPE as predictors. Separate between-group t-tests were calculated for choice probability and exploratory trials. Results were adjusted at the peak level for multiple comparisons using the family-wise error control. Small volume correction was performed using the following *a priori* regions of interests (ROIs): 1) the ventral striatum, using an anatomical definition of the nucleus accumbens (as obtained in the IBASPM atlas as part of the WFU Pick Atlas) with respect to SU and DU PEs; 2) the ventromedial prefrontal cortex (vmPFC) because of its central role in choice value, which is closely linked to DU PEs and choice probability. The vmPFC ROI was defined using a functional ROI of the effects of DU RPE and choice probability, respectively, published by a previous independent study on development of reversal learning (Waltmann, Herzog et al. 2023); 3) a functional ROI from the same previous study (Waltmann, Herzog et al. 2023) reflecting brain activation to noisy/ explorative behavior.

## Results

### Descriptive Statistics

17 ADHD patients and 17 age- and gender-matched healthy controls were included. Except for two left-handed ADHD patients, all participants were right-handed. One female participant was included in each group. According to the CIDI DIA-X screening interview, two subjects in each group fulfilled the diagnostic criteria of alcohol abuse (F10.1). Ten subjects in the ADHD group reported nicotine use, of which two subjects met criteria for nicotine dependence (F17.2). Four subjects in the control group reported nicotine use. In the ADHD group, two subjects had not previously been treated with stimulants, seven had been treated with methylphenidate in the past, and nine were still taking methylphenidate (but discontinued the medication 48 hours prior to the study appointment). As expected, compared with healthy controls, the ADHD group reported stronger ADHD symptom ratings in the CAARS and WURS-K questionnaires, but no differences in other psychiatric symptom ratings according to the Symptom Checklist (SCL-90) (Derogatis and Savitz 1999). Descriptive group statistics are presented in Table 1.

**Table 1.** Sample Description of Age, Clinical Symptoms and Handedness

|  | ADHD | HC |  |  |  |  |
| --- | --- | --- | --- | --- | --- | --- |
|  | *M ± SD* | *M ± SD* | *t-value* | *df* | *p* | *Effect Size* |
| Age (in years) | 22.14 ± 4.07 | 23.58 ± 3.47 | 189.000<sup>c</sup> |  | .131 | 0.31 |
| CAARS (t-score) |  |  |  |  |  |  |
| Inattention | 57.53 ± 12.25 | 46.25 ± 6.94<sup>a</sup> | 3.278<sup>b</sup> | 26 | .003* | 1.13 |
| Hyperactivity | 57.47 ± 8.52 | 42.94 ± 6.32<sup>a</sup> | 5.536 | 31 | <.001** | 1.94 |
| Impulsivity | 54.12 ± 10.59 | 42.56 ± 7.07<sup>a</sup> | 3.661 | 31 | .001** | 1.28 |
| Self-Concept | 52.76 ± 13.98 | 43.88 ± 5.03<sup>a</sup> | 2.457<sup>b</sup> | 20 | .023 | 0.85 |
| ADHD Index | 60.65 ± 11.77 | 43.13 ± 7.43<sup>a</sup> | 5.075 | 31 | <.001** | 1.78 |
| GSI t-score (SCL-90-R) | 53.06 ± 9.50 | 50.88 ± 7.33 | .748 | 32 | .460 | 0.26 |
| WURS-K (raw score) | 42.76 ± 14.93 | 22.18 ± 9.14 | 4.850 | 32 | <.001** | 1.66 |
| Handedness (raw score) | 32.06 ± 7.81 | 35.12 ± 1.58 | 1.584<sup>b</sup> | 17 | .131 | -0.54 |

*Note.* CAARS = Conners Adult ADHD Rating Scale (Conners et al., 1998), GSI= Global Severity Index, SCL-90-R = Symptom Checklist-90-R (Franke, 2002), WURS-K, Wender Utah Rating Scale' (Rösler et al., 2008). For the Student t-test and the Welch t-test, effect size is given by Cohen's *d*. For the Mann-Whitney U test effect size is given by the rank biserial correlation.
<sup>a</sup> *n* = 16. <sup>b</sup> Welch T-Test as equal variances were not assumed. <sup>c</sup> Mann-Whitney U-Test as not normally distributed. \* *p* < 0.01; \*\* *p* < 0.001

A detailed summary of the neuropsychological testing is presented in Table 2. The ADHD group had a lower intelligence in comparison to controls and performed worse than controls in working memory (digit span) and processing speed (TMT) domains.

**Table 2.** Neuropsychological Performance of ADHD Patients versus Healthy Controls

|  | ADHD | HC |  |  |  |  |
| --- | --- | --- | --- | --- | --- | --- |
|  | *M ± SD* | *M ± SD* | *t-value* | *df* | *p* | *d* |
| IQ (CFT-20-R) | 98.18 ± 15.96 | 108.82 ± 7.86 | 2.468<sup>a</sup> | 23 | .021* | -.85 |
| Digit Span |  |  |  |  |  |  |
| Forward | 6.65 ± 1.84 | 8.47 ± 2.00 | 2.767 | 32 | .009** | -.95 |
| Backward | 6.00 ± 1.73 | 7.53 ± 1.91 | 2.447 | 32 | .020* | -.84 |
| Total | 12.65 ± 3.26 | 16.06 ± 3.53 | 2.930 | 32 | .006** | -1.00 |
| TMT |  |  |  |  |  |  |
| Part A (in sec.) | 28.09 ± 5.16 | 23.61 ± 7.65 | 2.005 | 32 | .053 | .69 |
| Part B (in sec.) | 79.12 ± 20.92 | 60.12 ± 18.77 | 2.787 | 32 | .009** | .96 |
| Part B minus Part A | 51.02 ± 21.09 | 36.51 ± 16.39 | 2.240 | 32 | .032* | .82 |

*Note.* IQ = intelligence quotient, CFT-20-R = Culture Fair Test 20, revised (Weiß & Weiß, 2008), Digit Span (von Aster et al., 2006), TMT = Trail-making-test (Reitan, 1958). <sup>a</sup> Welch T-Test as equal variances were not assumed. \* $p < 0.05$ ; \*\* $p < 0.01$

*Behavorial Data*. Accuracy differed only marginally between phases (t = 1.75 (df = 7), p = 0.08) and not between groups (t = 0.08 (df = 7), p = 0.94). However, there was a significant group*phase interaction effect, as ADHD patients performed better in the post-reversal phase and worse in the pre-reversal phase (t = 4.7 (df = 7), p < 0.001, figure 1a).

**Figure 1:**
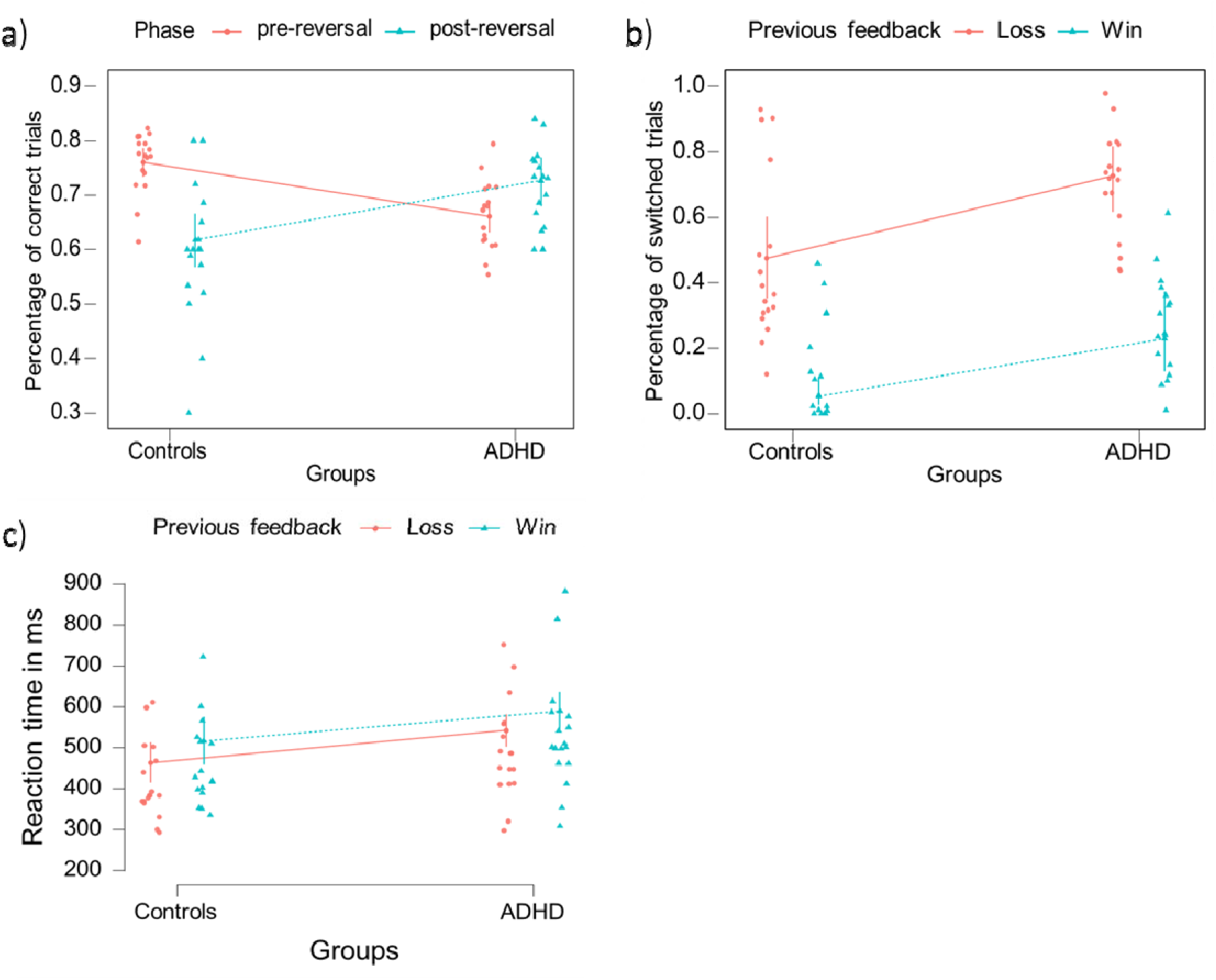
a) While controls chose the correct option more often during the stable pre-reversal phases, ADHD patients chose the correct answer more frequently during the post-reversal phase (Group*Phase interaction: t = 4.7 (df = 7), p < 0.001). Error bars depict standard errors of the mean. **b)** Subjects chose an option more often if it had been rewarded previously (feedback effect: t= 8.14 (df = 7), p < 0.001). ADHD patients were less likely to choose the same option twice in a row, regardless of the previous feedback (group effect: t= 4.12 (df = 7), p < 0.001). **c)** Both groups were faster to respond if they had lost in the previous trial (t = 4.1 (df = 7), p < 0.001). ADHD patients responded slower overall than controls (t = 2.3, p = 0.02).

All subjects were more likely to switch after previous negative feedback (feedback effect: t = 8.14 (df = 7), p < 0.001). ADHD patients were more likely to switch (group effect: t = 4.12 (df = 7), p < 0.001), irrespectively of previous feedback (group*feedback effect: t = 0.97 (df = 7), p = 0.33, see figure 1b).

All subjects responded faster when they had lost in the trial before (feedback effect: t = 4.1 (df = 8), p < 0.001, see figure 1c). ADHD patients responded more slowly than controls (group effect: t = -2.26 (df = 8), p = 0.02) independent of feedback (group*feedback effect: t = 0.23 (df = 8), p = 0.8).

### Computational Modeling of behavorial data

#### Model comparison

We compared a total of twelve RL models with respect to their evidence to account for the data based on the integrated Bayesian Information Criterion (Huys, Eshel et al. 2012). The double update model with separate learning rates and reinforcement sensitivities, as well as weighting of single and double updating, accounted best for the current behavioral data (see figure 2a).

**Figure 2:**
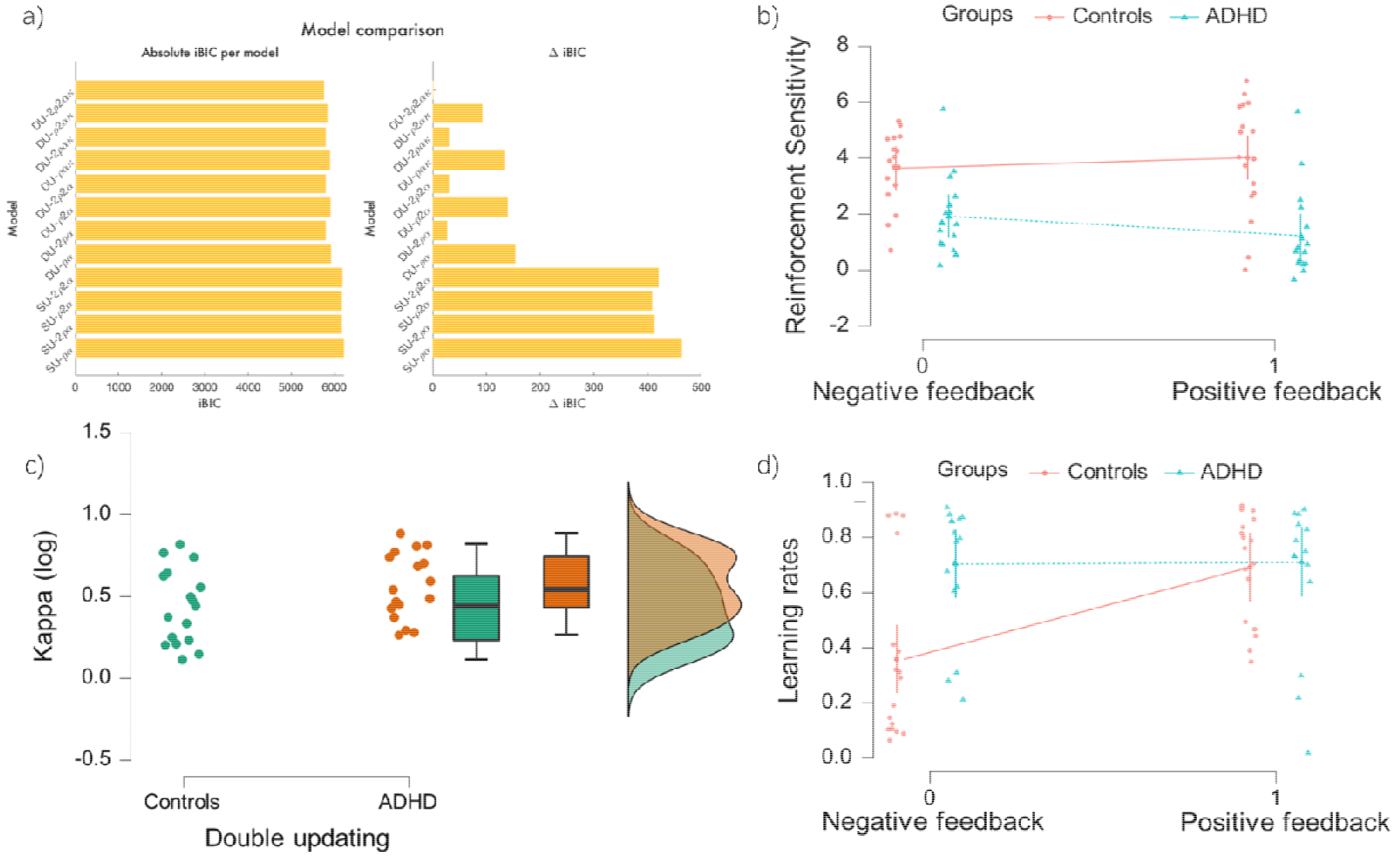
a) Model comparison showing the integrated Bayesian information criterion (iBIC) and delta iBIC (distance from the model with the best evidence) of the respective models. The most complex Q-learning model with separate learning rates and reinforcement sensitivities for wins and losses and a weighting parameter between single and double updating showed the best fit to the behavioral data. **b)** ADHD patients showed an overall lower reinforcement sensitivity (group effect: t = 4.3 (df = 6), p<0.001). Reinforcement sensitivity for positive feedback was especially lower in ADHD patients (group*feedback effect: t = 4.0 (df = 6), p<0.001). **c)** ADHD patients and healthy controls differed marginally in weighting the value of the chosen and non-chosen option to update, with ADHD patients updating the unchosen option slightly weaker. (double updating, p = 0.09, Cohen’s d = 0.59). **d)** ADHD patients showed a higher learning rate than controls (group effect: t = 2.5 (df = 6), p=0.02). This effect was driven by the higher learning rate for negative feedback in ADHD patients (group*feedback effect: t= 3.2 (df = 6), p=0.003).

#### Model parameters

ADHD patients showed an overall lower reinforcement sensitivity (group effect: t = 4.3 (df = 6), p < 0.001), especially for positive feedback (group*feedback effect: t = 4.0 (df = 6), p < 0.001), see figure 2b). The learning rate of ADHD patients was increased compared to healthy controls (group effect: t = 2.5 (df = 6), p=0.016), but this effect was mainly driven by the higher learning rate for negative feedback (group*feedback effect: t = 3.2 (df = 6), p = 0.003, see figure 2d). The parameter kappa, which defines the strength of the update weighting between chosen and unchosen option, showed a marginal difference between groups. ADHD patients updated the selected option slightly stronger than the unselected option compared to healthy controls (p = 0.09, Cohen’s d = 0.59, see figure 2c).

#### Correlations

In an exploratory analysis, which was not corrected for multiple comparisons, we correlated all five modeling parameters and the three core symptoms in ADHD patients (inattention, hyperactivity and impulsivity). Stronger hyperactivity symptoms (r=-0.5, p=0.04) and marginally stronger impulsivity symptoms (r=-0.42, p=0.09) were associated with a weaker updating of the unchosen option. Stronger impulsivity symptoms in ADHD patients were associated with lower learning rate for positive feedback (r=-0.51, p=0.03). Stronger hyperactivity symptoms were marginally associated with a lower learning rate for negative feedback (r=-0.5, p=0.07). Scatter plots of the significant correlations are shown in supplementary figures 1a and b. Exploratory analysis showed no further associations between modeling parameters and clinical symptoms (max r=0.27, min p=0.3).

#### fMRI analysis

Across both groups, single update prediction errors were significantly correlated with activity in the left and right ventral striatum (xyz: -13/8/-15, T: 3.51, p_FWE_: 0.003; xyz: 12/10/-15,, T: 2.55, p_FWE_: 0.045, see Figure 3a), but not in the ventromedial prefrontal cortex (xyz: 2/33/-2, T: 2.30, p_FWE_: 0.520). There were no significant differences between groups for the ROIs (VS L: xyz: -18/6/-15, T: 0.84, p_FWE_: 0.423; VS R: xyz: 14/3/-15, T: 1.23, p_FWE_: 0.342; VMPFC: VS L: xyz: -10/50/0, T: 2.74, p_FWE_: 0.277) nor on the whole brain level.

**Figure 3:**
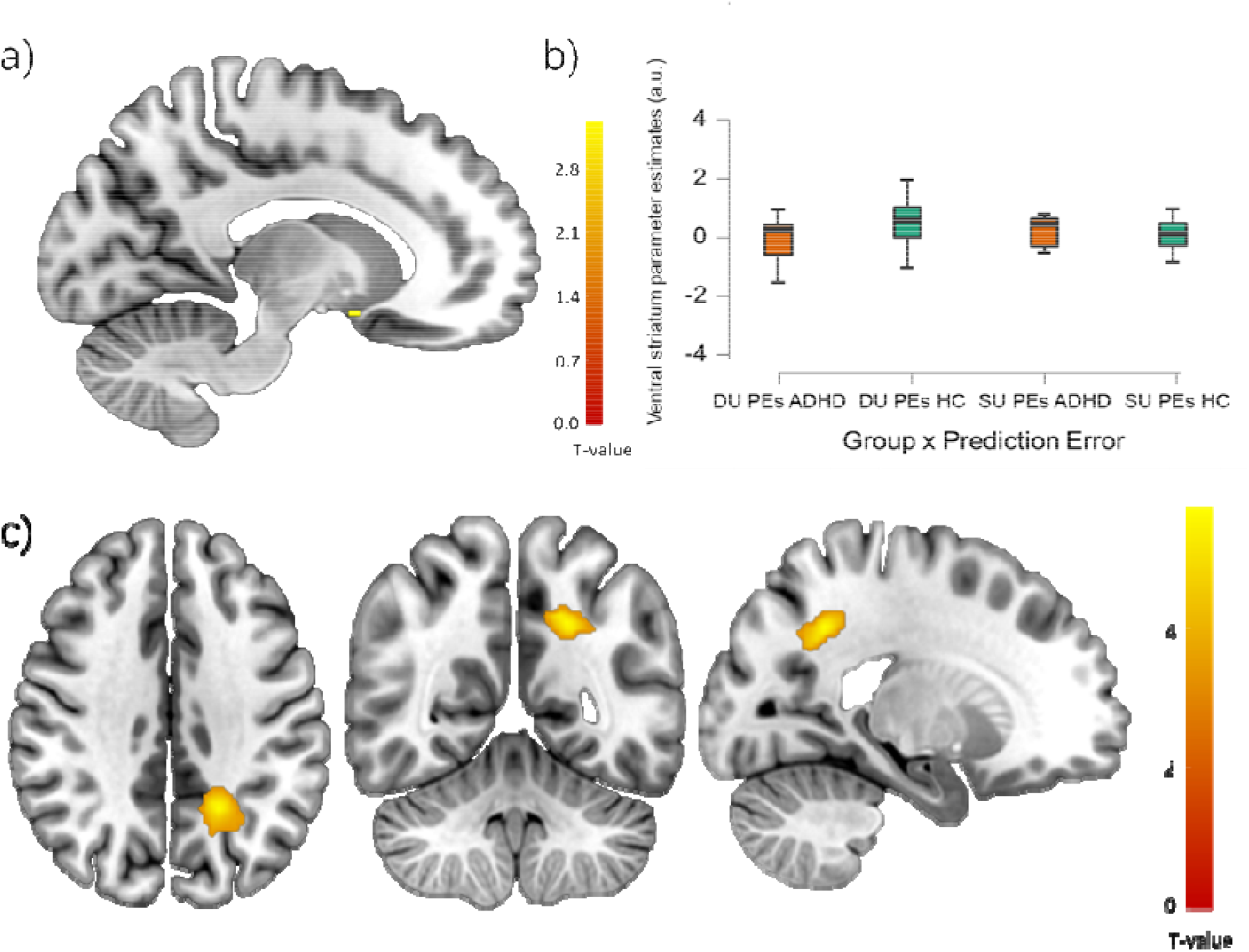
a) Single update prediction errors of both groups were represented in the left nucleus accumbens (small volume corrected for the ventral striatum: pFWE: 0.003, T: 3.51). **b)** There was a marginally significant prediction error x group interaction (PE x group interaction: T: 2.12, pFWE: 0.097). ADHD patients that had a weaker double update prediction error (DU PEs) signal in the right ventral striatum drove this effect. DU: Double update, SU: Single update, PEs: Prediction Errors, a. u.: Arbitrary units. **c)** Choice probability representation in the posterior parietal cortex was weaker in ADHD patients (pFWE: 0.04, T: 5.82, xyz: -20/-54/38). The color bars represent the t-values.

Across both groups, double update prediction errors were significant in the ventromedial prefrontal cortex cortex (xyz: 0/53/8, T: 3.62, p_FWE_: 0.043;) and only marginally significant in left (xyz: -13/10/-15, T: 2.31, p_FWE_: 0.060) and right ventral striatum (xyz: 7/6/-8, T: 2.37, p_FWE_: 0.066). We found a marginally higher double update prediction error signal in the right ventral striatum of healthy controls (type of PE x group, xyz: 7/6/-8, T: 2.12, p_FWE_: 0.097, see figure 3b) but not in the vmPFC (xyz: 0/6/-8, T: 2.95, p_FWE_: 0.191). No other group effects emerged on the whole brain level.

Across both groups, trial-by-trial choice probability was only marginally significantly related to activity in the vmPFC (xyz: 7/50/-8, T=3.66, pFWE=0.083). There was no group difference in the vmPFC (4/16/-8, T=2.16, pFWE=0.690). However, at the whole brain level, there was a significantly weaker neural representation of choice probability in ADHD as compared to controls in the posterior parietal cortex (p_FWE_: 0.04, T: 5.82, xyz: -20/-54/38, see figure 3c).

Across both groups, we found neural representations of noisy/exploratory trials in the left and right insular cortex in both groups at the whole brain level (left: p_FWE_: 0.001, T: 7.52, xyz: - 38/16/-12; right: p_FWE_: 0.024, T: 6.08, xyz: 40/28/-8). Using a ROI of activation in these trials covering the same regions from an independent study (Waltmann, Herzog et al. 2023), there was no group difference in these regions (xyz 40/30/-8, T=3.34, p_FWE_ =0.734).

## Discussion

In this study, adult ADHD patients showed impaired performance specifically when the learning environment was stable while performance was slightly improved after a reversal had occurred. This can be understood through an overall enhanced choice switching, which is maladaptive when the environment is stable but beneficial when changes occur. Our RL modelling explains this choice switching most clearly by a blunted sensitivity to positive and negative reinforcement. Additionally, an enhanced learning rate after negative feedback as well as a subtle tendency for reduced double-updating also contribute to elevated levels of choice switching. On the neural level, this was mirrored by a weaker representation of choice probability (which is scaled by reinforcement sensitivity) in the parietal cortex and weak indications for reduced double-update PEs in the right ventral striatum of ADHD patients. These results should be treated with caution, in particular with regard to double updating, due to the limited sample size of the current study.

A similar study in adolescent ADHD patients and healthy controls with a probabilistic reversal learning task in fMRI showed only partially overlapping results (Hauser, Iannaccone et al. 2014). This is probably partly due to different analysis methods such as the underlying models or the inclusion of pre- and post-reversal phases. The study also found no group difference in terms of overall accuracy, but did not test for possible phase effects, which we found to be significant. While modeling implementation was slightly different (in our modeling, we used a fixed softmax function and variable reinforcement sensitivities instead of variable temperatures of the softmax function) we find comparative results indicating enhanced decision noise leading to increased exploratory behavior.

As described above, individuals with ADHD had a significantly weaker neural representation of choice probability in the parietal cortex, compared to the control group. The parietal cortex is a crucial part of the attention network (Rushworth, Krams et al. 2001, Ptak 2012). A weaker attentional system might disrupt the processing of reinforcement information, making it difficult for an individual to accurately perceive and control the positive and negative consequences of their actions. This in turn might result in a reduced sensitivity to reinforcement, as seen in our modeling data, suggesting that ADHD patients probably need stronger reinforcements to update their choice values and to maintain certainty in decision making. A lower reinforcement sensitivity and a weaker processing of choice probability could lead to noisy/exploratory choice switching behavior independent of prior feedback, which was clearly evident in our behavioral data.

We did not replicate the weaker prediction error signals found in ADHD patients in the ventromedial prefrontal cortex in the previous study (Hauser, Iannaccone et al. 2014). Instead, we found a trend of weaker learning signals of the double update prediction error in the nucleus accumbens in ADHD patients. The decreased reinforcement sensitivity could make it more difficult for ADHD patients to build an internal model of contingencies. Therefore, they are more likely to respond to acute changes, which is beneficial post-reversal but detrimental pre-reversal. This is in line with solid evidence that ADHD patients prefer smaller immediate rewards for easier tasks as opposed to larger delayed rewards for more difficult tasks (Tripp and Alsop 2001, De Meyer, Beckers et al. 2019). For ADHD patients, this could also lead to poorer retrieval of internal choice values, resulting in more variability in reaction times (Kofler, Rapport et al. 2013, Véronneau-Veilleux, Robaey et al. 2022). We speculate that this decreased reinforcement sensitivity could be linked to our observation of weaker learning signals of the ventral striatum that incorporate chosen and unchosen action values. This finding could result from a reduced integration of environmental information (external sensory information or internal states) to learning signals due to aberrant dopaminergic neuromodulation. Research with animal subjects has already shown that dopaminergic neurons have a modulatory effect on neuronal and circuit flexibility, which ultimately leads to changes in behavior (Siju, De Backer et al. 2021). With further necessary empirical evidence, this could be regarded as an extension of existing ADHD dopamine theories (Tripp and Wickens 2008, Ziegler, Pedersen et al. 2016). In this regard, computational modelling is a helpful tool to further elucidate dopamine-based learning mechanisms in ADHD.

The literature regarding the learning rate of ADHD patients is not yet congruent. One theory proposes that the performance difficulties of ADHD patients in reward learning tasks may not be associated with deficits in learning, but with the sensitivity to reinforcements and the storing of cue-outcome contingences (Luman, Van Meel et al. 2009). This is supported by studies with tasks that explicitly test learning from losses and wins (Agay, Yechiam et al. 2010). However, in our data and model analyses, ADHD patients showed an increased learning rate for negative feedback, whereas the learning rate for positive feedback did not differ between groups. A higher learning rate for negative feedback would ensure that choice values are extinguished more quickly after a reversal and thus support enhanced switching (Ziegler, Pedersen et al. 2016) as seen in our data (but not in feedback-specific manner). Valence-dependent learning deficits have also been observed in a disease in which decreased cerebral dopamine concentrations play a role: Parkinson’s disease (PD). In one study, unmedicated PD patients learned less well from positive feedback compared to healthy controls, but this effect was reversed for negative feedback (Frank, Seeberger et al. 2004). The authors attribute this to the different direct (D1 receptor) and indirect (D2 receptor) pathways of the basal ganglia: While reduced phasic dopamine bursts would decrease sensitivity to positive feedback (via D1 receptors), reduced tonic dopamine could provide increased D2 receptor activity supporting learning from negative feedback. According to this theory, influences of the tonic dopamine concentration and thus on the indirect pathway would influence the learning behavior of ADHD patients. Lower tonic dopamine levels and thus higher D2 receptor activity could thus enhance learning from negative feedback and in addition to augmented decision noise lead to increased switching behavior. While this would explain our data, our study cannot prove this theory. Further animal studies, for example by influencing tonic and phasic dopamine bursts by genetic manipulation (Beeler, Daw et al. 2010) are necessary to draw clearer conclusions.

### Limitations

The sample size of this study, n=17 per group is too small to draw more than preliminary conclusions. Caution is necessary also because small sample sizes can inflate effect sizes (Button, Ioannidis et al. 2013). While we matched the two groups for age, handedness, and gender, group differences emerged with respect to nicotine use, intelligence, and working memory. While it is known that ADHD patients are more likely to smoke (Ilbegi, Groenman et al. 2018) and perform worse on tests of intelligence (Bridgett and Walker 2006) and working memory (Kofler, Singh et al. 2020), these differences between groups may explain some of the behavioral differences. However, we believe that our binary choice task places relatively low demands on working memory. Nevertheless, one should be cautious in interpreting the results as generalizing to all ADHD patients. Finally, the question arises to what extent the task structure can really represent exploratory behavior. While there is some uncertainty in the currently used task, the strictly anti-correlated structure scarcely represents the real complex exploration behavior of ADHD patients in their everyday life.

## Conclusion

Using computational reinforcement learning models, this study provides insight into the neurocognitive processes that facilitate behavioral differences in motivational learning and decision making in ADHD patients. Noisier behavior of ADHD patients was associated with decreased reinforcement sensitivity in our study. This behavior could be due to a reduced neural representation of dopaminergic prediction error signals in the nucleus accumbens and a reduced representation of choice probability in the posterior parietal cortex in ADHD patients. We speculate that lower tonic dopamine levels might lead to faster relearning after negative feedback via D2 receptor activation in ADHD, which may prove beneficial in rapidly changing environments.

## Supporting information

Supplementary plots 1 a+b

## Data Availability

All data produced in the present study are available upon reasonable request to the authors.

