## Supplementary plots 1 a+b for "Impaired flexible reward learning is associated with blunted reinforcement sensitivity and attenuated learning and choice signals in ventral striatum and parietal cortex of ADHD patients"

Supplementary Figure 1a

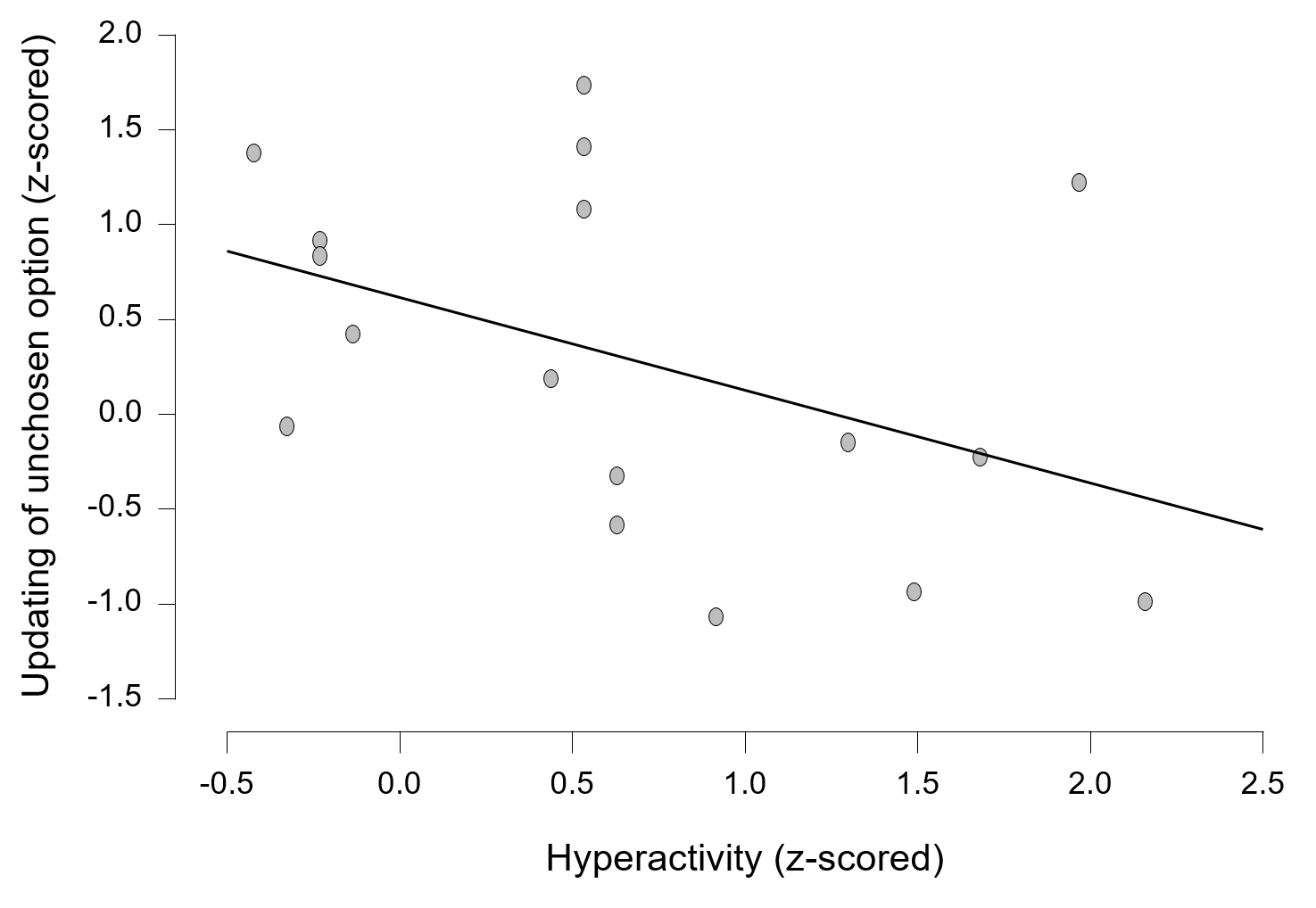

The more hyperactive the ADHD patients were, the less they updated the value of the unchosen action (r=-0.5, p=0.038).

Supplementary Figure 1b

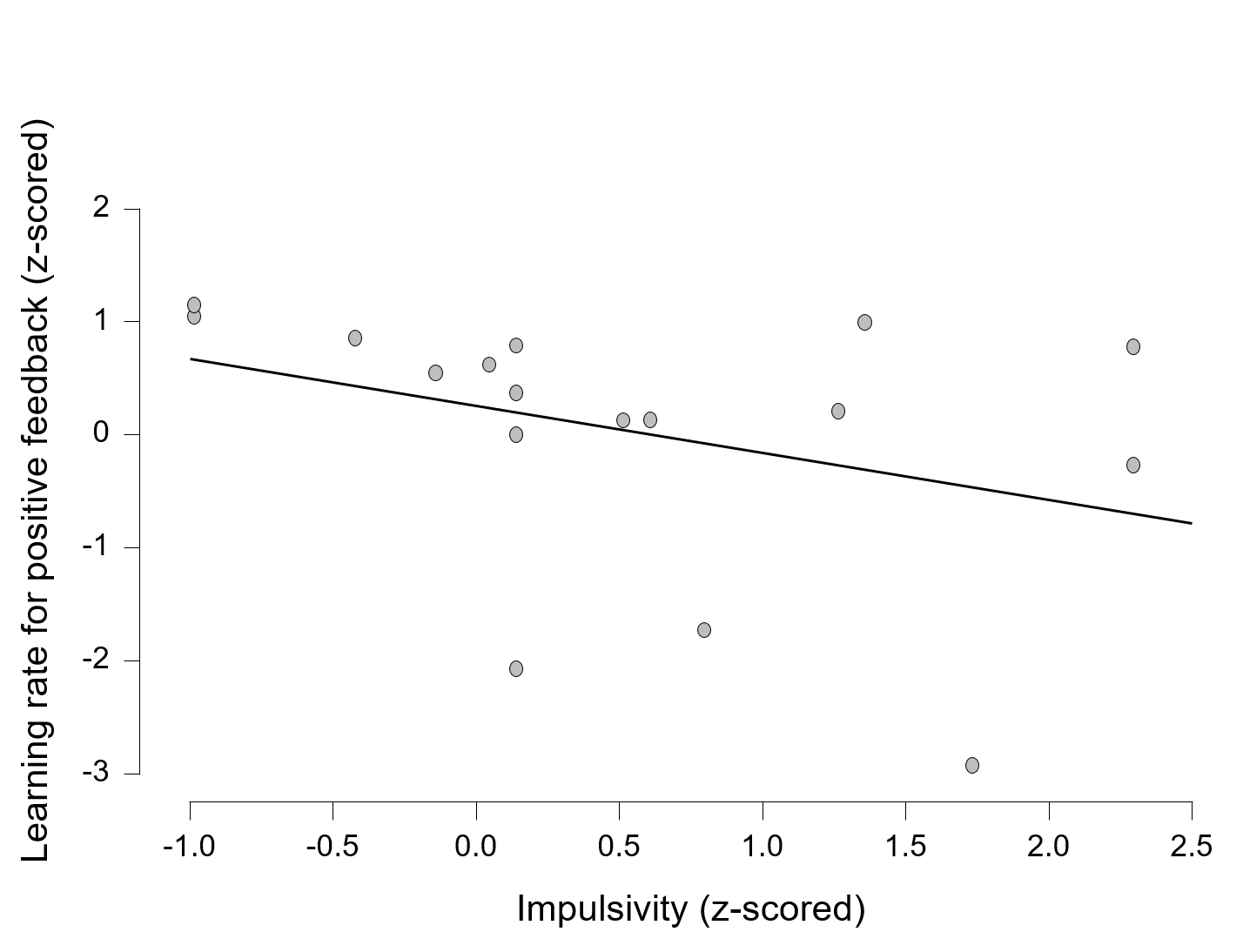

The more impulsive the ADHD patients were, the lower their rate of learning positive feedback (r=-0.5, p=0.036).
